# DEVELOPING A PROTOCOL USING WEARABLE CAMERAS FOR MEMORY TRAINING WITH OLDER ADULTS: A METHODOLOGICAL REPORT

**DOI:** 10.1101/2025.11.17.25339784

**Authors:** Tuíla Maciel Felinto, Tory Worth, Erin Welch, Roberto Cabeza

**Affiliations:** Universidade Federal do Rio Grande do Sul, Avenida Paulo Gama, 110, 90040-060, Porto Alegre, Rio Grande do Sul, Brazil; Duke University, 308 Research Dr, Durham, NC 27710; Columbia University, 1180 Amsterdam Ave, New York, NY 10027

**Keywords:** Autobiographical memory, wearable cameras, methodological report, older adults

## Abstract

Although not as pronounced as in Alzheimer’s Disease, healthy aging is associated with substantial deficits in memory functions, particularly in episodic and autobiographical memory. Efforts to train these impaired functions are hindered by the lack of transfer of the training effects to other tasks and daily activities and limited by artificial laboratory stimuli. Addressing these limitations, we developed an autobiographical memory training protocol using wearable cameras to record participants’ own life events as photos, and laptops to train their memory using these events at home. We tested the feasibility of the protocol in a study with 15 healthy older adults. The results showed that the protocol and set of instructions we designed enabled participants to use the equipment (wearable camera and laptop) successfully at home. In a novel addition, we used automated image processing to protect the privacy of the participants’ photos, which are considered Protected Health Information. Older adults’ responses to the overall study and camera use was positive, indicating that studies using wearable cameras can be engaging and motivating, and potentially even successful in improving memory. The methods will be the central focus on this paper, and the results will be expanded upon in a subsequent paper.

## Introduction

Even in healthy aging, older adults typically present cognitive deficits compared to younger adults, and the specific nature of these deficits continue to be investigated. Some studies show that processing speed, attention, and memory performance decline with age (Harada et al., 2013), although there is emerging conflicting evidence. Specifically, memory deficits have been associated with reduced quality of life and independence (Montejo et al., 2012) and are a common complaint among older adults (Reid & MacLullich, 2006). Autobiographical memory (memory for events naturally occurring in daily life) is a combination of episodic memory (memory for individual events and their associated contextual details; such as memory for a particular birthday party) and semantic memory (acontextual memory for repeated and generalized events, such as “playing tennis in college”). The episodic memory component is more impaired in healthy aging than the semantic component of autobiographical memory, particularly memory for the contextual details of individual events (Levine et al., 2002). This deficit can have serious implications in older adult life, such as misremembering that a medication was taken today when it was actually taken yesterday. Therefore, it is important to research how to improve memory performance or reduce cognitive deficits in older adults.

Evidence shows an association between cognitively stimulating activities performed throughout life and reduced age-related cognitive decline (Krivanek et al., 2021). Memory training interventions are a common type of training targeted at older adults that usually focus on teaching mnemonic strategies to be employed by participants in their daily lives (Allé et al., 2017). These interventions can be brief and focused on a single technique, or extended, including practices of different strategies, both in group or individual settings (Rebok et al., 2007).

Reviews show that different types of cognitive and memory training interventions can be effective in improving the specific ability for which they were trained, but the results usually cannot be generalized to or have minimal effects on other cognitive functions and everyday activities (Bahar-Fuchs et al., 2019; Bahk & Choi, 2018; Ball et al., 2002). Typically, memory training interventions focus on practicing mnemonic strategies such as rehearsal, association, categorization, imagery, concentration, or a combination of those (Gross et al., 2012). The participants learn how to use these strategies in their daily lives to help them to better remember specific information. These strategies are only useful tools in situations in which the person knows they are going to need to remember specific information later, which includes only a fraction of situations in real life.

One alternative to these kinds of memory training is to use everyday life events in tasks that aim to stimulate autobiographical memory, making the training more similar to the way memory naturally functions. However, it is difficult to gather enough life events from one person to compose a full intervention. One alternative solution to this issue is using wearable cameras to collect photos from people’s daily lives. Wearable cameras are small devices that can be worn in front of the body and take photos automatically from a first-person perspective. They are a simple and low effort approach to gathering a personalized stimuli set that can be used to help stimulate episodic memory while also allowing for accurate performance assessment (Hodges et al., 2011). Compared to traditional methods of memory research and rehabilitation that use laboratory stimuli (lists of generic images or words, for example), wearable cameras provide meaningful and personalized photos that can be used as stimuli in autobiographical memory tasks, allowing for greater ecological validity. In addition, the fact that photos are taken automatically, without the user’s intervention, allows events to be encoded incidentally and naturally, as it happens when the camera is not being used. The photos’ first-person perspective is closer to how autobiographical memories are encoded, making them efficient recall cues. Also, the personalized nature of the stimuli makes the task more interesting, relevant, and engaging (Allé et al., 2017).

The first studies investigating the use of wearable cameras were case studies evaluating the effect of reviewing the photos collected during selected events in the memory performance of patients with memory impairment. Berry et al. (2007) compared the use of wearable cameras to a written diary as a memory aid in a case study of a patient with limbic encephalitis; a condition that decreased her ability to remember remote and recent past experiences. The results showed that reviewing the images from a wearable camera improved the participant’s ability to recall past events and was considered more interesting and motivating than using a written diary. Other studies comparing wearable cameras with written diaries also found that photos from wearable cameras are more efficient in improving autobiographical memory and that participants find using the device more interesting (Browne et al., 2011; Woodberry et al., 2015). Wearable cameras allow the user to have a set of photos taken from a first-person perspective without interference, which is especially useful when working with patients with memory impairment who would find it difficult to remember to take photos during their daily activities (Berry et al., 2007). Additionally, if participants are asked to actively take the photos during a study, it is difficult to determine whether their performance on a memory task is due to increased attention during encoding or to the study’s manipulation.

Despite their advantages, wearable cameras do present some methodological challenges. The wearable camera takes photos automatically, according to a predetermined time interval; because of that, by the end of the day, the large volume of data collected can be an issue. Additionally, the randomly taken photos are occasionally blurred or too dark, which can undermine the effectiveness of an intervention using this method. One solution - tasking the research team to filter and organize this data into meaningful personal events - presents a host of privacy issues. Since the camera is lightweight and discrete, it is possible for participants to forget that they are wearing the camera and the photos collected might contain sensitive information. This issue can be avoided by asking the participants to review their own photos prior to sharing them with the research team, deleting the ones that they are not comfortable with sharing. This approach was reported in a study by Harvey et al. (2016). Their investigation combined a wearable camera and an activity monitor to examine how well older adults could use technology to observe sedentary behavior. The participants were six community-dwelling older adults who wore the camera for seven consecutive days. They were instructed to ask permission from others to wear the camera in private spaces and were informed in advance that they could ask for any of their photos to be deleted. When the participants returned to the lab after wearing the camera, they reviewed and deleted their photos before the researcher had access to them. A set of images from one participant took approximately 60 minutes for the researcher to review. The participants reported that the equipment was easy to wear and had little effect on their daily lives, although they reported functional difficulties. They also felt the camera did not affect their privacy, probably due to the ability to choose whether to share their photos or not. While this approach avoided privacy issues, it was very time-consuming, and it would be complicated to execute with a larger sample.

Using images collected automatically from people’s homes can lead to serious privacy issues that involve the participant, their family and friends, and even people in public spaces. Hoyle et al. (2014) focused on investigating the privacy concerns people have when using lifelogging technology. The authors asked 36 undergraduates to wear a smartphone with a lifelogging application for five days. The application was programmed to collect data from the smartphone’s accelerometer, magnetometer, and GPS, to take photos from 8 am to 10 pm, and to send the data to the research team via Wi-Fi. The participants could choose to pause the data collection or delete photos for a specific period while wearing the device. After each day, they would review their images and determine which should be deleted, flag blurred images to be deleted later, and label images by place. Also, the participants were asked to report why they chose to pause the app or delete photos during the day and explain which ones they were comfortable sharing. The authors found out that the factors involved in photosensitivity included location, people, and personal information visible in monitors or phone screens. They concluded that wearable devices should allow people to pause and delete images while using them, and that the participants must be instructed to only wear the device in situations in which actively taking photos is allowed, in order to protect the privacy of other people not involved in the study.

This methodological report is part of a larger project that aims to develop a memory training protocol that uses relevant stimuli specific to each participant while avoiding interference in their daily lives. To achieve this goal, we developed a training protocol in which older adults take home a wearable camera and a laptop computer and perform the training by themselves. We aimed to formulate a protocol structure that would allow the participants enough time to learn how to deal with the equipment and the tasks involved in the training. This study describes four pilot studies of this protocol, each building upon the previous one’s strengths while improving specific aspects based on the participants’ feedback. Also, based on the results of previous studies that used wearable cameras, we intended to reduce the privacy and technical issues reported, using image processing software that evaluates photo quality (filters out blurred images) and groups the photos into events by similarity without human intervention (Doherty & Smeaton, 2008). This procedure allows for reduced time and effort in reviewing photographs from a large sample and avoiding privacy issues. The photos are only presented on the screen during the memory task, and the participants are the only ones to see them. All sensitive information that can be present in the images was already available to that person when the photo was taken, thus avoiding the need to review and remove images before presenting them.

## Method

### Participants

This paper describes four pilot studies, each building upon the previous one according to the researcher’s observations and feedback from the participants. The pilot studies had 3, 2, 6, and 4 participants, respectively. The final sample included 15 participants (8 women) between 64 and 80 years old (M = 72, S D = 4.92). The participants were recruited through fliers posted around Duke University campus and at public community spaces nearby. The study was conducted under an approved Duke Health IRB protocol with Roberto Cabeza as its principal investigator.

### Materials

Each participant received a wearable camera (Narrative Clip 2) and a laptop. The Narrative Clip 2 is a wearable lifelogging camera measuring 3cm x 3cm x 1.5cm and weighing 19g. The camera takes photos at a 3264 x 2448 pixels resolution, and it can store up to 4000 images onboard (8GB memory). The images can be downloaded to a computer using a USB cable via the Narrative Uploader (version 1.7.0.0). This proprietary software downloads the photos in the camera to the computer and starts automatically when the Narrative Clip is connected. The camera comes with a clip and a lanyard to be worn either attached to the clothes or around the neck. According to tests performed by our laboratory staff, the lanyard is the best option because it gives the camera greater stability, reducing the number of blurred photos. The camera was initially programmed to take one photo every two minutes to reduce download and image processing times, but this was changed to one per minute for the final pilot study.

The laptop was a Dell Inspiron 14 5000 Series 2-in-1 5481 with an Intel i3 processor, 8GB RAM, and a 256GB SSD storage drive running on Windows 10 Home. Before the beginning of the study, each laptop was prepared to avoid any interruptions from the operational system. All pre-installed apps were uninstalled, all system notifications were hidden, and all automatic updates were blocked. The system updates and the overall state of the computers were checked at the laboratory before the beginning of the study. We installed two software applications: the Narrative Uploader and the Memory Task program. The Memory Task program was developed by the researcher in Python v3.8.8. It integrates the image processing software (the Quality Identifier and the Segmentation software) developed by consultants Alan Smeaton, Cathal Gurrin, and their team from Dublin City University (Doherty & Smeaton, 2008). The first software evaluates the quality of the photos, calculating values for features such as darkness and definition, labeling them as *clear* or *blurred*. The threshold for the selection can be adjusted, and after several tests, we decided on a threshold of 0.15. The second software parses the set of clear images into events, by comparing each image with the following ones and calculating the similarity between their visual features. According to our tests, comparing each image to the following three allowed for a more accurate representation of real-life events. The Memory Task program is responsible for integrating the image processing software and presenting the memory task using the remaining clear photos grouped into events. The images were stored in the computer, and the image processing was done locally, which increased the safety of the participant’s sensitive data.

The participants were instructed on how to use the camera and the laptop during the first laboratory visit and took home a detailed instructions sheet. The final version of the instructions sheet included screenshots from the computer, step-by-step information on how to use the computer and start the program, and what to do if one of the programs did not start. It also contained the researcher’s contact information, and the participants were instructed to call or email if they had issues with the equipment.

### Procedures

The participants received detailed explanations about the study, and the researchers answered all questions about privacy and safety regarding the use of the camera. After that, the participants who agreed to take part in the study signed the consent form. More detailed instructions detailing how to use the camera and what to do in specific situations were provided during the appropriate time. The structure and duration of each pilot study varied according to the participants’ feedback and the researchers’ observations. These differences and the reasoning behind all changes are discussed in the Results section.

The first pilot study had only one phase (training phase) and two lab visits. In contrast, the other three were divided into two phases (familiarization and training phase) and three lab visits (Figure 1). During the familiarization phase, the participants only wore the camera and downloaded their photos on the computer every day at home. In contrast, during the training phase, they additionally performed a memory task. In this task, they were presented with two events (short clips that included five photos from their camera) and would choose which one was the most recent by touching the image on the screen. In the third and final pilot studies, the participants were tested at the lab before and after training by doing the task with events not used during training. Therefore, another camera-only phase (similar to the familiarization phase) was added to collect photos for the last memory task at the lab. The memory task results are not reported or discussed in this paper.

**Figure 1.**
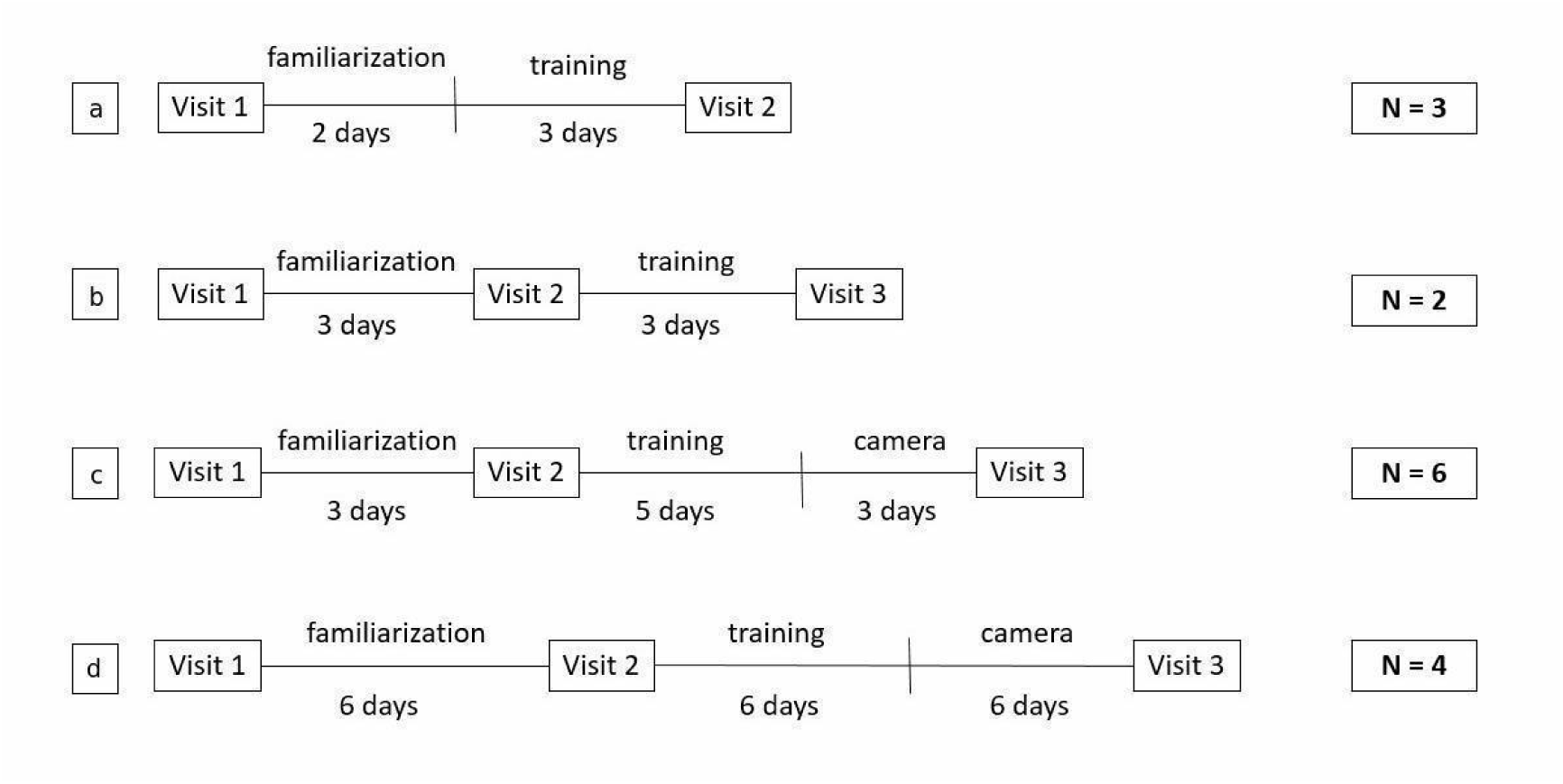
Timeline of the four different pilots of the memory training study. In pilot one (a), the first visit included both equipment and task instructions. In the remaining pilots (b, c, and d), the first visit included equipment instructions and the second visit included task instructions.

After each pilot, the instructions sheet was improved, and more details were added based on the participants’ feedback to make sure that they had all the information they needed to use the equipment at home by themselves. The participants could call or email the lab if they had any problems with the equipment or questions about the procedures. The participants were asked to wear the camera for around six hours per day. They could choose when and where to wear it, but they were instructed to prioritize periods when they would be more active. Since the camera does not have an off button (like many other wearable cameras available), anytime the participant felt that it was not appropriate to use a camera, they could turn it around to have its lens covered or take it off and put it into a pocket.

At the end of the day, the participants downloaded the photos from the camera and started the Memory Task program. The procedure was simplified, so even participants who had no familiarity with technology could use the equipment at home. After connecting the wearable camera to the computer with an USB cable, the Narrative Uploader software would start automatically and download the photos from that day. Then, participants clicked the Memory Task program icon on the desktop and waited. The Memory Task program then started both the Quality Measure software, which deleted blurred photos, and the Segmentation software, which grouped the remaining photos into events by comparing each image with the following three (Doherty & Smeaton, 2008). Then, depending on the phase, the program would present the memory task or not. To help participants remember the instructions, the desktop wallpaper had an arrow indicating where the window of the camera software would appear after photos were downloaded, and an arrow pointing to the Memory Task icon that participants should click once the download was finished.

### Data analysis

The participants’ feedback and a summary of the calls and emails received during the study were compiled from the researcher’s notes. This data was interpreted and reported in the results when relevant to explain the development of the protocol. The count of useful images (total of photos taken in a day minus blurred or dark images) and events in each pilot study was analyzed to index how well the participants could use the camera (Gelonch et al., 2019). Also, the image processing time was reported as a measure of the efficiency of the procedure.

## Results

### Procedure development

The first pilot study included two visits and a five-day phase divided into two days of familiarization and three training days (Figure 1a). On the first visit, participants were instructed on how to wear the camera, download the photos from that day, and do the memory task. During the familiarization phase, the participants only wore the camera and downloaded the images. During the training phase, they also did the memory task with the photos collected. One of the participants had issues dealing with the computer on the second day after wearing the camera successfully on the first day. It was not possible to solve the problem over the phone, so the participant did not complete the study. The other two participants were able to complete the procedure, but they reported during the last visit that they were not sure how to answer the memory task. These results indicated that the large number of instructions provided during the first visit were confusing since the participants were not used to the technology and were already worried about learning how to deal with the wearable camera.

The second pilot study aimed to reduce the amount of information given on the first visit (Figure 1b), which focused only on explaining how to use the camera and download the photos for the familiarization period. After that, they returned to the lab and were instructed on how to do the memory task. In this version, they were only presented with an example of the task on the screen during the second visit. All the participants were able to complete the study, and all issues with the camera were solved via email or over the phone. Some participants reported feeling a little confused about the task initially but understood it better after some trials at home.

The third pilot study also had three visits and two phases, but the training phase included a photo collection period before the last visit (Figure 1c). The participants were tested at the lab before and after the training. The events presented at the lab test during the second visit were collected during the familiarization phase. The events presented at the lab test during the last visit were collected during this period after the training, making sure they were all new events. In addition to that, during the second visit, the participants were presented with a few example trials using their photos from the familiarization phase so they could better understand how the task worked. In all of the previous pilot studies, most of the participants were initially confused about the computer touchscreen: they would first try to answer the task by pressing the computer keys and then they would remember that they should touch the computer screen. Although most of our participants were at least somewhat used to the technology (they were contacted via email and all had smartphones), only a few were familiar with computer touchscreens. Using their own photos in the example trials was useful in helping them overcome this issue because it helped them to better associate the demonstration with the actual task compared to using events from someone else’s life as an example.

By the end of the training, participants complained about finding it difficult to identify when some of their events happened because the images represented things they frequently did. Because of this issue, we changed the frequency of the photos and the way events were composed in the final pilot study (Figure 1d). The wearable cameras were programmed to take one photo per minute instead of a photo every two minutes to provide more detailed events and to account for events that were too similar. The images are grouped into events automatically by the Segmentation software, and they can be parsed into groups of different lengths. In the first three pilots, the first five images of each event were presented (Figure 2a). In this pilot, however, there was a more holistic approach. The first photo of the event, the last picture, and the middle photo were presented to the participant, as well as the photos that were halfway between the first photo and the middle picture, and the middle photo and the last photo (Figure 2b). These changes allowed for more diverse events making the difference between the events in a trial more clear and the memory task more interesting for the participants.

**Figure 2.**
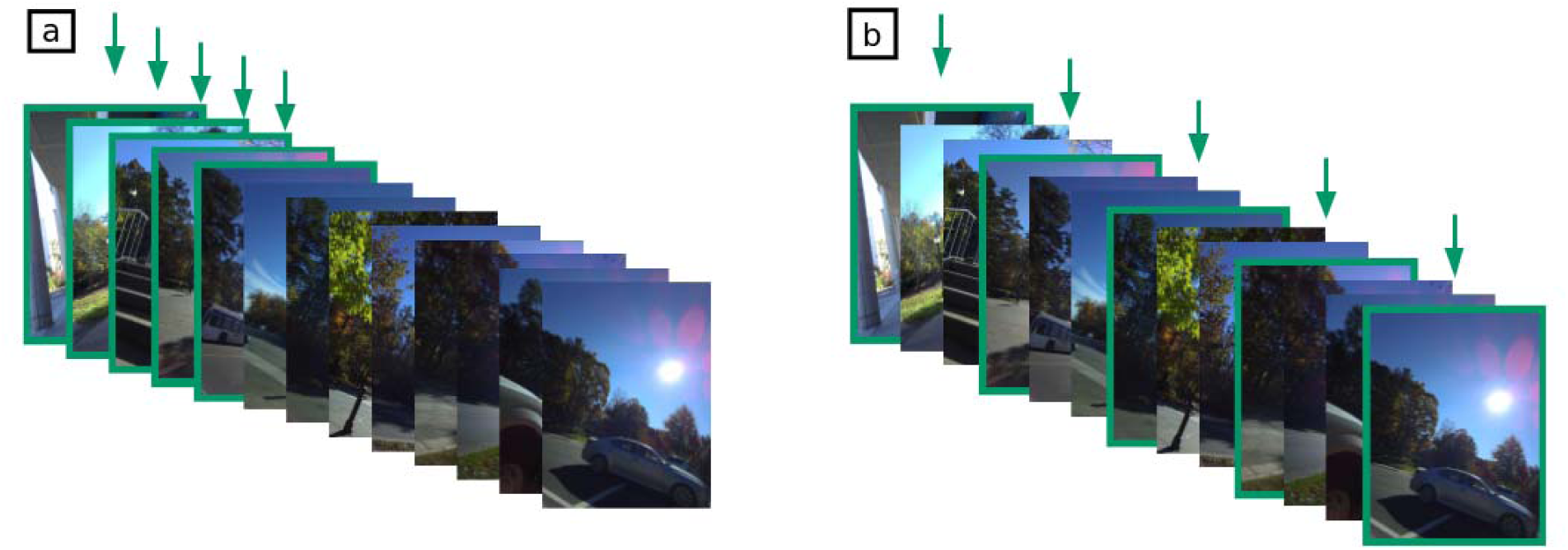
Image selection for event composition in the first three pilots (a) and in the final one (b).

### Instructions and participant feedback

Considering the participant feedback from our final pilot study, the study structure and instructions were clear enough to allow an easy adaptation to the equipment and comprehension of the task. This final pilot included detailed information on basic actions such as how to turn the computer on and off, how to log in to the computer, and where to type the computer password. By the last pilot, we divided the instructions sheet into two parts, which were given to the participant during the first and second lab visits. The first part included information on dealing with the computer and the camera, and how to download photos and start the Memory Task program to process them. The second part reviewed information about the training task specific for each group. During the second visit, the task was explained, and the participants did an example version and a full version to get baseline data. Even with this previous experience, we noticed that some participants needed to review the instructions at home before starting the program and doing the task by themselves.

This instruction division allowed the participants to ask more detailed questions during the visits and have a clearer understanding of what they should do at home since they had less information to handle at each stage. The participants’ feedback during all pilots allowed us to notice the aspects of computer handling that this specific population finds challenging. During pilot three, one of the participants emailed us to ask how to get the laptop to open. After this episode, we made sure to ask how familiar the participant was with that specific kind of equipment and clearly demonstrated every step of the process unless they stated they knew how to do it. Some older adults may be used to handling specific devices (their own smartphone or computer) but still get confused about aspects that are new to them (the computer model, a different operating system).

### Camera issues and data

This study used the Narrative Clip 2, a small and discrete lifelogging camera that is simple to use. Although the Narrative Clip 2 has these advantages and captures high-quality images, our participants encountered some issues while wearing it during their day-to-day activities. When the camera is connected to the computer via USB cable, the Narrative Uploader software is supposed to launch automatically and start downloading the photos. The most common complaint from participants was that sometimes the Narrative Uploader software did not launch by itself. After the second pilot study, we included a shortcut for the Narrative Uploader on the computer’s desktop. We instructed the participants to double-click the icon in case the software did not launch after some time. Also, the participants complained that the back of the Narrative Clip 2 could get very hot while it is charging. Those specific cameras presented no issues to capture and download the photos, which leads to thinking that the temperature while charging is not a problem.

The percentage of useful photos in each pilot indicates that the participants were able to wear the camera correctly, especially after the second pilot, even with the difficulties encountered. After the changes were applied to the last pilot, the mean number of events increased, favoring a more detailed representation of the participants’ daily lives (Table 1). The image processing times, which includes detecting and deleting blurred photos (QM duration) and grouping the remaining images into events (SEG duration), was kept below ten minutes, even when the frequency of image capture was increased to one per minute (Table 2).

**Table 1.** Summary of images collected, useful images, and useful events per day.

| Version | Valid | Missing | Pictures |  | Useful pictures |  |  | Useful events |  |
| --- | --- | --- | --- | --- | --- | --- | --- | --- | --- |
|  |  |  | *M* | *SD* | *M* | *SD* | % | *M* | *SD* |
| 1 | 11 | 0 | 252.45 | 176.51 | 141.36 | 104.46 | 55.99 | 7.09 | 4.86 |
| 2 | 11 | 0 | 189.36 | 68.59 | 139.9 | 55.64 | 73.88 | 6.81 | 3.15 |
| 3 | 47 | 0 | 149.42 | 78.61 | 96.51 | 52.67 | 61.24 | 5.23 | 3.03 |
| 4 | 54 | 0 | 433.44 | 210.07 | 260.94 | 130.21 | 60.2 | 12.64 | 7.39 |
| Total | 123 | 0 | 291.29 | 208.6 | 176.59 | 124.15 | 60.62 | 8.79 | 6.48 |

**Table 2.**
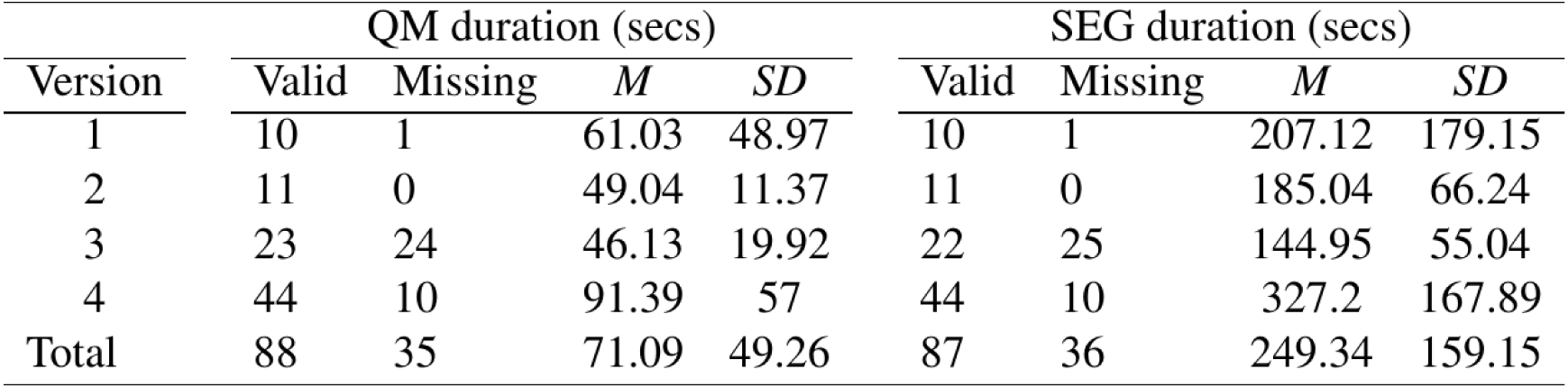
Summary of quality and segmentation processing times.

## Discussion

This study aimed to develop a memory training protocol that older adults could do at home with remote support from professionals. We described how the protocol was gradually improved after each pilot study in order to adapt to the participants’ needs. In the past decade, there has been an increase in the number of studies that translate cognitive training aimed at older adults to computers, tablets, and smartphones. These tools allow the researchers to monitor a larger number of participants in real time and remotely. Also, it is possible to adapt the frequency of tasks and the difficulty level to each participant’s performance (Ge et al., 2018; Kueider et al., 2012; Lampit et al., 2014). Using different technological tools to develop training protocols enables the exploration of other types of stimuli previously inaccessible through traditional research methods. Personal and recent photos, videos, and audio recordings, and the combination of these types of media can be used to investigate the mechanisms of cognitive processes and how they work outside the laboratory setting. Results from studies show that images from wearable cameras are effective as autobiographical memory cues and that reviewing these images led to more recall of details that were not visible in the photos (Mair et al., 2018). Other studies found that interventions involving the reviewing of wearable camera images had an effect on global measures of autobiographical memory (Silva et al., 2013, 2017).

### Privacy

This protocol aimed to minimize issues raised by previous studies (Allé et al., 2017; Kelly et al. 2013). Collecting photos automatically from a first-person perspective can lead to recording the potentially sensitive visual information of both the study participants and third parties (family, coworkers, and strangers). Previous studies accounted for these issues by including a review session when participants could delete any photos they wanted to before allowing the research team to access the images. This procedure is time-consuming and can be a problem in studies in which the photos would be used as stimuli in memory tasks. In such cases, the review session could interfere with the study manipulation. Regarding those aspects, Kelly et al. (2013) proposed a framework for ethical practice in studies using wearable cameras. The authors addressed issues concerning the research participants’ privacy and third parties that may be unknowingly photographed during the study. They argue that no image should be shared, and no one should be identified in the photos without consent. The authors also highlight that, since participants may forget they are wearing the camera after some time, they should be allowed to review the images and delete those they are not comfortable sharing with the research team. Furthermore, the participants should be clearly informed of what to do in situations where someone asks questions about the camera (contact the research team), how the images will be stored, and legal problems that may arise (in case the research team sees something illegal happening in one of the photos). The sensitive nature of the material being collected requires careful assessment of all stages of the study to guarantee that the research team is prepared to handle it and that the participant’s privacy is not being violated.

Our protocol accounted for this issue by ensuring that the participant would be the only person to see the images. The photos are only presented during the memory task. They are stored locally, and all necessary image processing is done automatically. The software developed by our consultants Alan Smeaton, Cathal Gurrin, and their team (Doherty & Smeaton, 2008) detected blurred images and segmented the remaining ones into events by image similarity. Then, our Memory Task program used the segmented events to compose the memory task, present it, and collect the data. The research team only had access to the participant’s task performance data and the program log files. These files were monitored to assure that the equipment was working and the participant was using the camera correctly (this information could be inferred by checking the number of photos collected per day, the number of images deleted, and the number of events generated). After the last visit, the Memory Task program automatically deleted all photos stored since the beginning of the study.

During the instructions session on the first visit, the participants were instructed about what to do if someone else felt uncomfortable with the camera: they should give them the laboratory’s contact information and take the camera off (Kelly et al., 2013). The participants reported that only close friends and family asked about the camera and that, after a brief explanation about the study, no one expressed any additional concerns. Our lab never received any calls or emails from third parties regarding the study. We believe that the Narrative Clip 2, as are other wearable cameras, is small and discreet enough to not draw much attention from strangers in situations where it would be acceptable for someone to use their smartphone to take photos.

During the instruction sessions, one should consider the specific resources and limitations of each participant (Schmidt & Wahl, 2019) since some may benefit from more detailed information or reviews. During the first visit, the participants learned how to use the camera and the computer in an individual instructions session. Every part of the procedure was demonstrated, including basic steps such as turning on the computer. Our participants ranged from significantly familiar with technology (e.g., a retired computer engineer) to minimal familiarity (the participant could use their personal computer but got confused with the laptop assigned by the laboratory). Therefore, the research team decided that it was more effective to ask questions during the instruction session to assess the participant’s understanding and adapt the amount of details provided according to each participant.

The participants seemed more confident in their ability to complete the study correctly when they were informed that they could contact a researcher if they had any problems or questions at any point during the study. All participants contacted the laboratory via email or phone at least once during the study. Aside from one occurrence (participant from pilot one, see the Results session of this paper), all issues were solved. While most of them preferred to talk over the phone, they had no problems contacting the laboratory via email. Maintaining open communication with the participants allowed them to call with questions or issues that could lead to losing data if left unaccounted. The participants were informed that the study aimed to make the procedure easier for them to follow and that made them more comfortable expressing their thoughts, questions, and complaints about the equipment and the instructions.

During the visits, the participants were asked for feedback about the equipment, instructions, and procedures. They reported specific points about the instructions sheet that they believed were missing or could use more details. All suggestions were incorporated, and the participants from the final pilot reported that the instructions sheet helped them remember details about the camera and the tasks. We received positive feedback about the overall experience of the study during all pilot studies. Some participants reported that they enjoyed reviewing photos from their past days during the memory tasks. Others said that, although they felt self-conscious at first while wearing the camera, the feeling did not last long, and they soon forgot they were wearing it. Similarly, in a study from Gelonch et al. (2019), participants reported they believed that the benefits of the wearable camera outweighed the perceived inconveniences. In our study, one participant asked where they could purchase a similar camera for personal use and another asked for reading material about the study. The feedback supports the results from previous wearable camera studies that compared the review of wearable camera photos and written diary entries (Berry et al., 2007; Browne et al., 2011; Woodberry et al., 2015).

## Conclusion

Our results show that our final visits’ structure and set of instructions allowed the participants to understand how to use the equipment correctly at home by themselves - 60% of the photos taken in a day were clear and useful, in line with findings in Gelonch et al. (2019). Furthermore, our participants’ responses to the overall study and camera use were positive, indicating that studies using the wearable camera can be engaging and motivating. An important advance in the current study is the automatization of the image processing stage which allowed for increased protection of the participants’ privacy compared to previous studies (Harvey et al., 2016; Hoyle et al., 2014). The process of filtering poor quality images and segmenting the remaining ones into events was done locally and automatically without the need for either the participant’s or the researcher’s manual review, reducing the amount of time and effort needed for selecting events for the task. Moreover, monitoring the participants’ progress during the training phase through log files generated by the Memory Task program made it easier to monitor multiple participants simultaneously and reduced the need for remotely accessing the participant’s computers to solve specific problems.

During the pilots described in this study, the participants reported that they sometimes found it difficult to temporally localize events depicting activities they do frequently, such as driving, working on the computer, or watching television. While decreasing the interval between photos from one photo every two minutes to one photo per minute led to more detailed events, the problem was attenuated, but not solved. Future research could deal with this issue by including a software module that identifies repeated events and selects the most distinct ones to be used in the task (Doherty et al., 2011). Another issue that occurred at a much lower rate was that participants reported seeing blurred photos during the task. Increasing the sensitivity of the clearness threshold of the Quality Measure software could solve this problem, but it would also lead to more clear photos being classified as blurred. A possible solution for this problem would be adding a way for the participants to flag poor quality images that would then be removed from the tasks.

Finally, it is important to note that the camera used during this study is a commercial product, not developed specifically for research use and not adapted to older adults, which led to issues concerning the structure of the procedure. The photos collected by the Narrative Clip 2 can only be accessed via the Narrative Uploader software which adds an extra step in the participant’s daily task. Studies in autobiographical memory using personal images would benefit from a camera developed with research needs in mind, such as flexibility and transparency.

## Declarations

## Data Availability

All data produced in the present work is contained in the manuscript. Code is available at the URLs provided.

https://www.dropbox.com/s/j7ab2bqo8v4csme/QM.zip?dl=0

https://www.dropbox.com/sh/pf32qdnngwlknld/AABoxo0v3nqPKznCTr9rUgKGa?dl=0

## Acknowledgements and funding

This project was supported by the National Institutes of Health (NIH) grant number 1-R21-AG059006-01. The authors acknowledge the contributions of Cathal Gurrin, Alan Smeaton, and collaborators in developing the image processing and segmenting software used in this study.

## Ethics approval

This study was approved by Duke University IRB (Pro00100512).

## Consent to participate

Informed consent was obtained from all participants included in the study.

## Consent for publication

Not applicable.

## Availability of data and materials

All relevant data are provided in the content of the manuscript. This study was not preregistered.

## Code availability

The code for the image quality measurement (QM) and image segmentation (SEG) software are available at the URLs below.

QM: https://www.dropbox.com/s/j7ab2bqo8v4csme/QM.zip?dl=0

SEG: https://www.dropbox.com/sh/pf32qdnngwlknld/AABoxo0v3nqPKznCTr9rUgKGa?dl=0

